## Supplementary Information for "The health impacts of a 4-month long community-wide COVID-19 lockdown: Findings from a prospective longitudinal study in the state of Victoria, Australia"

**Supplementary Table S1. Summary of restrictions leading to, during, and following the 2020 extended community lockdown in Victoria, Australia.**

| Restrictions | Date | Location in Victoria (VIC) |
|---|---|---|
| Postcode lockdown | 30 Jun | 10 postcodes areas in Melbourne |
| Postcode lockdown | 4 Jul | 2 additional postcodes areas in Melbourne |
| Interstate border closed between New South Wales (NSW) and Victoria | 8 Jul | NSW-VIC border |
| **Stage 3 restrictions.**<br>Four reasons to leave home:<br>• Shopping for essential items<br>• Caregiving<br>• Exercise (1 hour)<br>• Work (employers must support you to work from home if you can work from home) | 8 Jul | Metropolitan Melbourne and Mitchell shire |
| Schools return to flexible and remote learning for pupils in Prep to Year 10 (i.e. ages 5/6 to 15/16) | 20 Jul | Metropolitan Melbourne and Mitchell shire |
| Face coverings mandatory | 22 Jul | Metropolitan Melbourne and Mitchell Shire |
| Visitor limits. No hosting visitors at home or visiting people at their home | 30 Jul | Otway-Colac region (6 local government areas west of Melbourne) |
| **Stage 4 restrictions.**<br>• Curfew from 8pm to 5am<br>• 5km distance limit from home.<br>• 1-hour exercise limit<br>• Maximum gathering of two.<br>• Shopping limited to one person per household.<br>• Weddings banned (from 5 Aug)<br>• All onsite students learning from home (5 Aug) | 2 Aug | Metropolitan Melbourne |
| Face coverings mandatory | 2 Aug | VIC |

| | | |
|---|---|---|
| **Stage 3 restrictions.**<br>Four reasons to leave home:<br>• Shopping for essential items<br>• Caregiving<br>• Exercise (1 hour)<br>• Work (employers must support you to work from home if you can work from home) | 5 Aug | Regional Victoria (including Mitchell Shire) |
| **Easing restrictions:** | | |
| **Step 1:**<br>**Metro Melbourne:**<br>Curfew from 9pm to 5am<br>One permitted visitor (bubble) for people living alone and single parents<br>Exercise and social interaction (2 hours)<br>Increased gathering limits<br>Onsite learning staged return from Term 4 (PREP to Grade 2, VCE and VCAL 12 Oct)<br><br>**Step 2**<br>**Regional VIC:**<br>One permitted visitor (bubble) for people living alone<br>Gatherings limit: 5 people outdoors from 2 households<br>Outdoor public pools and playgrounds open<br>Religious services outdoors: limit 5 people and a faith leader<br>Onsite learning staged return from Term 4 (all students between 12-16 Oct) | 13 Sep | Metropolitan Melbourne and Regional Victoria |
| **Melbourne:**<br>5km limit to 25 km limit<br>Any reason allowed for leaving home<br>No time limit on exercise and social interaction<br>Outdoor sport settings reopen<br>Allied health professionals resume face-to-face care<br>Hairdressers reopen<br>Gatherings limit: 10 people from 2 households in outdoor public places<br><br>**Regional VIC:**<br>Libraries limit of 20 people indoors<br>Outdoor religious gatherings limit 20 people<br>Hospitality: 40 customers indoors, up to 70 outdoors | 18 Oct | Metropolitan Melbourne and Regional Victoria |
| **Regional VIC + Greater Shepparton**<br>Gyms and fitness spaces: up to 20 people, density 1 person per 8 square metres<br>Indoor pool limit: 20 people<br>Indoor sport begins for people aged 18 and under<br>Food courts open<br>Live music resumes (outdoor hospitality) | 27 Oct | Regional VIC and Greater Shepparton |

| | | |
|---|---|---|
| Religious celebrations: 20 people limit indoors, 50 people outdoors, and a faith leader | | |
| **112 days of lockdown ends**<br>**Metro Melbourne**<br>Further relaxation on visitors<br>Any reason permitted to leave home<br>25km limit from home<br>Ring of steal in place (work permits required for crossing boundary)<br>ring of steal in place<br>Restaurants and pubs reopen: limit of 50, 20 indoors<br>Remaining retail opens<br>Workplaces reopen ("if you can work from home you must work from home" in place) | 27 Oct | Metropolitan Melbourne |
| **Step 3**<br>No restrictions on intrastate travel<br>Restrictions eased in Metropolitan Melbourne to be consistent with regional VIC<br>2 people visit per day (different households, different visits) | 8 Nov | VIC |
| 15 visitors each day<br>Outdoor gathering limit 50<br>Religious ceremonies limit 150 people indoors<br>Hospitality: density limit 1 person per 2 square metres, up to 50 customers. QR code record keeping mandatory. Larger venue cap of 150 people. | 22 Nov | VIC |
| Masked limited to: public transport, some retail and indoor shopping centres / stores, supermarkets.<br>30 people visitor limit<br>Outdoor gatherings limit 100<br>Density limit restaurants, café, pubs 1 person per square metre, no cap. QR code record keeping mandatory. | 6 Dec | VIC |

**Supplementary Table S2. Comparisons of participants included in the analyses (Retention group) and those excluded (Attrition group).**

| Group N (%) | **Retention group**<br>Completed 3 surveys at all relevant time-points (baseline, 3- and 6-month follow-up surveys). | **Attrition group**<br>Completed baseline survey, but did not complete at least one of the 3- or 6-month follow-up surveys |
|---|---|---|
| **Total** | **898 (100.0)** | **1705 (100.0)** |
| **Demographics** | | |
| ***Gender**** | | |
| Female | 475 (52.9) | 1139 (66.8) |
| Male | 421 (46.9) | 554 (32.5) |
| ***Age group*** | | |
| 18 to 24 years | 67 (7.5) | 178 (10.4) |
| 25 to 34 years | 123 (13.7) | 319 (18.7) |
| 35 to 44 years | 150 (16.7) | 334 (19.6) |
| 45 to 54 years | 218 (24.3) | 429 (25.2) |
| 55 to 64 years | 268 (29.8) | 384 (22.5) |
| 65 or more years | 72 (8.0) | 61 (19.6) |
| **Pre-existing health** | | |
| ***Anxiety*** | | |
| Yes | 106 (11.8) | 357 (20.9) |
| No | 792 (88.2) | 1348 (79.1) |
| ***Depression*** | | |
| Yes | 139 (15.5) | 353 (20.7) |
| No | 759 (84.5) | 1352 (79.3) |
| **Survey mode** | | |
| Online form | 186 (20.7) | 1037 (60.8) |
| Telephone interview | 712 (79.3) | 668 (39.2) |

*aggregate data with group sizes < 10 are not disclosed due to ethical considerations.

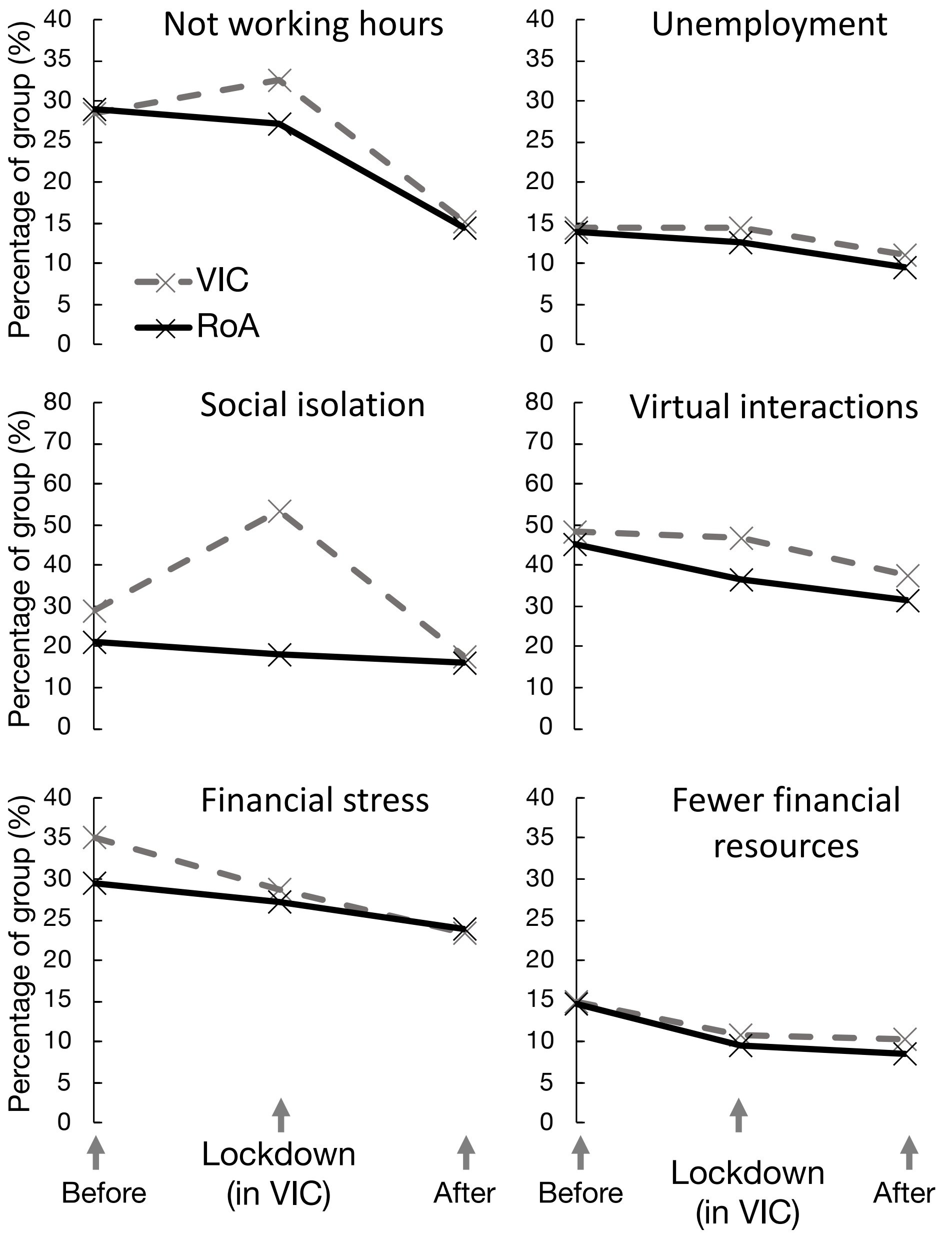


**Supplementary Figure S1. Changes in determinants of health prior to, during, and after the community lockdown in Victoria, compared to the Rest of Australia.**

Note: VIC – the Australian state of Victoria (i.e. lockdown location during 8 July – 27 October 2020). RoA – Rest of Australia.

**Supplementary Table S3. Impacts of the lockdown in Victoria on determinants of health including work, social interactions and finance.**

| | Adjusted Odds Ratios [95% Confidence Interval] | | | | | |
|---|---|---|---|---|---|---|
| | **Work** | | **Social interactions** | | **Finance** | |
| | ***Not working hours*** | ***Unemployment*** | ***Social isolation*** | ***Virtual interactions*** | ***Fewer financial resources*** | ***Financial stress*** |
| **The Lockdown Impact** | | | | | | |
| During lockdown | **2.00** [1.40, 2.87] | 1.17 [0.77, 1.77] | **3.73** [2.52, 5.53] | 1.33 [0.96, 1.85] | 1.15 [0.73, 1.79] | 0.83 [0.59, 1.16] |
| After lockdown | 1.08 [0.69, 1.68] | 1.13 [0.67, 1.91] | 0.75 [0.49, 1.15] | 1.15 [0.82, 1.64] | 1.28 [0.81, 2.01] | 0.74 [0.52, 1.07] |
| **Changes in health over location and time** | | | | | | |
| VIC * pre-lockdown | **2.83** [1.96, 4.09] | **1.75** [1.13, 2.71] | **2.20** [1.57, 3.08] | **2.10** [1.57, 2.82] | **1.85** [1.17, 2.92] | **1.76** [1.28, 2.42] |
| VIC * lockdown | **2.58** [1.74, 3.80] | **1.73** [1.11, 2.68] | **6.49** [4.68, 9.00] | **1.95** [1.45, 2.62] | 1.24 [0.76, 2.05] | 1.26 [0.90, 1.77] |
| VIC * post-lockdown | 1.14 [0.73, 1.78] | 1.27 [0.79, 2.04] | 1.16 [0.80, 1.68] | 1.31 [0.97, 1.75] | 1.19 [0.72, 1.98] | 0.95 [0.67, 1.35] |
| RoA * (pre-lockdown) | **2.68** [2.01, 3.58] | **1.56** [1.13, 2.16] | **1.43** [1.10, 1.85] | **1.86** [1.51, 2.28] | **1.98** [1.49, 2.64] | **1.37** [1.11, 1.70] |
| RoA * (lockdown) | 1.22 [0.95, 1.56] | **1.32** [1.03, 1.69] | 1.13 [0.88, 1.45] | **1.30** [1.05, 1.60] | 1.16 [0.85, 1.59] | 1.20 [0.97, 1.47] |
| RoA * (post-lockdown) | 1.00 (ref.) | 1.00 (ref.) | 1.00 (ref.) | 1.00 (ref.) | 1.00 (ref.) | 1.00 (ref.) |

Estimates with $P < .05$ shown in bold. VIC – the state of Victoria (i.e. lockdown location during 8 July – 27 October 2020). RoA – Rest of Australia. Models were adjusted for gender, age group and survey mode. *The Lockdown Impact* describes health differences of working-age Victorians to the Rest of Australia, controlling for health differences pre-lockdown.

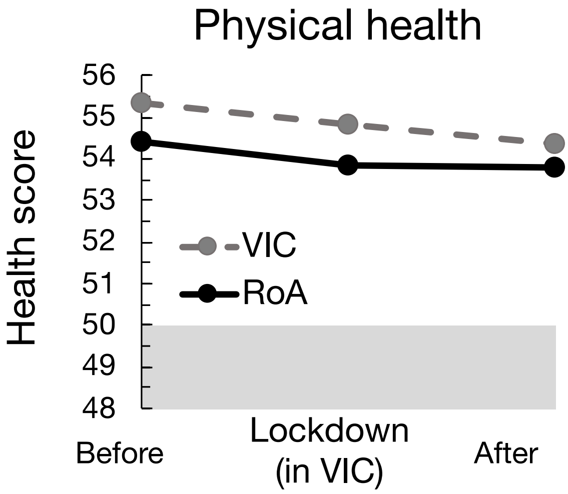


**Supplementary Figure S2. Changes in average physical health scores prior to, during, and after the community lockdown in Victoria compared to the Rest of Australia.**

Note: Shaded regions indicate moderate-high distress or below (pre-pandemic) average health; VIC – the Australian state of Victoria (i.e. lockdown location during 8 July – 27 October 2020). RoA – Rest of Australia.

**Appendix A. Media statements and Victorian Government publications on restrictions (corresponding to those listed in Supplemental Table S1).**

Andrews D. Statement From The Premier. [media release] (2020 July 7) [cited 2021 June 3]. Available from https://www.premier.vic.gov.au/statement-premier-74

Andrews D. Statement From The Premier. [media release] (2020 June 30) [cited 2021 June 3]. Available from https://www.premier.vic.gov.au/statement-premier-72

Andrews D. Statement From The Premier. [media release] (2020 July 4) [cited 2021 June 3]. Available from https://www.premier.vic.gov.au/statement-premier-73

Berejiklian G. Border closure to protect NSW. [media release] (2020 July 6) [cited 2021 June 3]. Available from https://www.nsw.gov.au/media-releases/border-closure-to-protect-nsw

Andrews D. Return To Flexible And Remote Learning. [media release] (2020 July 12) [cited 2021 June 3]. Available from https://www.premier.vic.gov.au/return-flexible-and-remote-learning

Andrews D. Face Coverings Mandatory For Melbourne And Mitchell Shire. [media release] (2020 July 19) [cited 2021 June 3]. Available from https://www.premier.vic.gov.au/face-coverings-mandatory-melbourne-and-mitchell-shire

Andrews D. Statement From The Premier. [media release] (2020 July 30) [cited 2021 June 3]. Available from https://www.premier.vic.gov.au/statement-premier-75

Andrews D. Statement On Changes To Melbourne's Restrictions. [media release] (2020 August 2) [cited 2021 June 3]. Available from https://www.premier.vic.gov.au/statement-changes-melbournes-restrictions

Andrews D. Statement On Changes To Regional Restrictions. [media release] (2020 August 2) [cited 2021 June 3]. Available from https://www.premier.vic.gov.au/statement-changes-regional-restrictions

Andrews D. Statement From The Premier. [media release] (2020 September 6) [cited 2021 June 3]. Available from https://www.premier.vic.gov.au/sites/default/files/2020-09/200906%20-%20Statement%20From%20The%20Premier.pdf

Victoria State Government. Metropolitan Melbourne – Summary of easing of restrictions at 11:59pm on Sunday 18 October and Sunday 1 November. Oct 2020. https://www.premier.vic.gov.au/sites/default/files/2020-10/201018_Metro_Melb_.pdf

Victoria State Government. Regional Victoria – Summary of easing of restrictions at 11:59pm on Sunday 18 October and Sunday 1 November. Oct 2020. https://www.premier.vic.gov.au/sites/default/files/2020-10/Regional%20VIC_Easing%20of%20Restrictions_.pdf

Andrews D. Statement from the premier. [media release] (2020 October 18) [cited 2021 June 3]. Available from https://www.premier.vic.gov.au/statement-premier-77

Victoria State Government. Regional Victoria - Summary of further easing in the Third Step. Oct 2020. https://www.premier.vic.gov.au/sites/default/files/2020-10/Regional%20VIC%20Easing%20Restrictions%2025-10%20V3_converted.pdf

Victoria State Government. Metro Melbourne - Summary of the Third Step and further easing of restrictions. Oct 2020. https://www.premier.vic.gov.au/sites/default/files/2020-10/201026%20-%20Metro%20Melb%20Easing%20Restrictions.pdf

Victoria State Government. Summary of statewide restrictions for the Third Step and Last Step of Victoria's roadmap to reopening. 2020 Nov. https://www.premier.vic.gov.au/sites/default/files/2020-11/201108%20-%20Third%20Steps.pdf

Victoria State Government. Summary of Last Step Restrictions from 11.59pm On 22 November 2020. 2020 Nov. https://www.premier.vic.gov.au/sites/default/files/2020-11/221120%20-%20Last%20Step%20restrictions%20.pdf

Victoria State Government. COVIDSafe Summer – How we work in Victoria. 2020 Dec 6. https://www.premier.vic.gov.au/sites/default/files/2020-12/201206%20-%20COVIDSafe%20Summer%20-%20How%20we%20work.pdf

Victoria State Government. COVIDSafe Summer – How we live in Victoria. 2020 Dec 6. https://www.premier.vic.gov.au/sites/default/files/2020-12/201206%20-%20COVIDSafe%20Summer%20-%20How%20we%20live.pdf
